# Developing Predictive Algorithms for Patient Retention Using Machine Learning and Deep Learning to Improve HIV Care in Uganda

**DOI:** 10.1101/2025.11.20.25340654

**Authors:** Alex Mirugwe, Solomon Ssevvume, Arthur G. Fitzmaurice, Jonathan Mpango, Alice Namale, Evelyn Akello, Paul Katongole, Simon Muhumuza, Naseef Mayanja, Paul Mbaka, Enos Sande, Kenneth Musenge

## Abstract

**Background:** Achieving high retention of people living with HIV (PLHIV) in care remains a challenge in Uganda, despite substantial progress towards UNAIDS 95-95-95 targets. This study used advanced machine learning and deep learning techniques applied to de-identified longitudinal PLHIV data routinely collected in HIV clinics in Uganda to predict clients at high risk of missing treatment appointments.

**Methods:** We compared the performance of traditional machine learning models (i.e., Decision Tree, Random Forest, AdaBoost, and XGBoost) and the Bidirectional Encoder Representations from Transformers (BERT) model, which is more suitable for analyzing longitudinal data. Feature importance using the Shapley additive exPlanations method was used to identify the most influential predictors. We also evaluated the impact of various sampling techniques, i.e., undersampling, oversampling, and synthetic minority oversampling, to address class imbalance and improve model performance. Model performance was evaluated using accuracy, precision, recall, F1-score, and the Area Under the Curve-Receiver Operating Characteristic (AUC-ROC) metrics.

**Results:** The study was based on a longitudinal dataset of 66,206 PLHIV who initiated HIV care during 2000-2023 in 86 health facilities. The data comprised 1,479,121 clinical visits, an average of 22 clinical visits; 158,266 (10.7%) missed appointments, and 49,588 (74.9%) of clients missed at least one appointment. Median (interquartile range [IQR]) age was 36.0 [29.0 – 47.0] years, and the majority (n=43132, 65%) were female. The BERT model demonstrated superior performance, achieving an AUC score of 0.96, 94.8% accuracy, 97.1% precision, 100% recall, and an F1-score of 94.2%. In comparison, the XGBoost model with undersampling achieved an AUC score of 0.90, 80.7% accuracy, 97.1% precision, 80.8% recall, and an F1-score of 88.2%. Feature importance analysis showed that treatment adherence, visit frequency, treatment duration, and visits on the current regimen were the most influential predictors of appointment interruption.

**Conclusion:** This study highlights the efficacy of transformer-based models like BERT in handling longitudinal clinical data and improving patient retention predictions. Integrating these predictive models into electronic medical systems will facilitate proactive treatment strategies, enabling the identification of clients at risk of disengagement before they miss appointments. This approach may contribute to the improvement of HIV care and support progress towards achieving the HIV program targets in Uganda and potentially elsewhere.

## 1 Introduction

Despite significant progress towards achieving the UNAIDS 95-95-95 targets in Uganda, where 94% of people living with HIV (PLHIV) know their HIV status, 92% are on ART, and 84% are virally suppressed, HIV/AIDS remains one of the leading causes of death, with AIDS-related deaths standing at approximately 22,000 annually [1]. A major challenge still persists in consistently retaining PLHIV in care, which ultimately impacts the third 95 target. According to recent studies, 21% of PLHIV get lost within the first twelve months after antiretroviral therapy (ART) initiation, with variations across different regions [3]. Other studies report attrition rates ranging from approximately 10% to 25% [4–6]. High attrition rates in HIV medical services have been identified as a significant barrier to optimal HIV care, contributing to poor viral load suppression and, in turn, increased HIV transmission [7– 9].

The current strategies for re-initiation of lost to follow-up PLHIV require the use of community health workers (CHWs) and phone call reminders. The use of CHWs is expensive and often challenged by issues of patient confidentiality. Phone calls are often hindered by the lack of technological infrastructure, which is very common in many low-middle-income countries (LMICs). It is therefore important that active measures be put in place that can identify PLHIV who have a potential for loss to follow-up; these can benefit from health education and other targeted services to enhance their retention in care.

Recent studies have demonstrated the potential benefit of machine learning in predicting patient retention in HIV care [10–14]. Maskew et al. [10] applied machine learning to predict patient retention and viral load suppression in South African HIV treatment cohorts. Using algorithms like logistic regression and random forest on demographic and clinical data, they achieved AUCs of 0.69 for visit attendance and 0.76 for viral suppression. Ramachandran et al. [11] developed a model using electronic medical records and geospatial data to predict retention in an urban HIV clinic in Chicago, outperforming traditional methods with a positive predictive value of 34.6% for the top 10% of highest-risk patients. In another study on Pre-Exposure Prophylaxis (PrEP) retention among key populations in urban Zimbabwe, logistic regression and clustering techniques were used to identify significant predictors such as population type, sex, marital status, employment type, age, and education level, with female sex workers demonstrating the highest retention rates [15]. These studies highlight the potential of predictive modeling to enhance targeted interventions in HIV care and prevention programs.

Despite these advances, all these studies used traditional machine learning models to analyze longitudinal treatment data. While effective for many tasks, these models generally treat records as independent and are less able to capture sequential dependencies [16, 17]. Transformer-based models, particularly Bidirectional Encoder Representations from Transformers (BERT), are more suitable for handling sequential data due to their ability to capture contextual information from both directions and learn complex patterns [18, 19]. In addition, prior studies often relied on highly imbalanced datasets without applying sampling techniques, which likely biased their models toward the majority class, even when reporting good performance scores. Against this background, we propose utilizing machine learning algorithms to predict and identify PLHIV at high risk of missing their treatment appointment schedules. By identifying these PLHIV before they get lost to follow-up, we can enable early and tailored interventions, thereby improving retention in care. Therefore, this study aims to address the following key questions: i) How do traditional machine learning models compare to the BERT model in predicting missed appointment treatment dates among HIV patients using longitudinal clinical data? Ii) What is the impact of different sampling techniques on the performance of traditional machine learning models when faced with an unbalanced dataset? iii) Which combination of model and sampling technique yields the best performance across various evaluation metrics, and what factors contribute to this outcome?

## 2 Material and Methods

### 2.1 Study Design

This was a retrospective design that utilized quantitative machine learning and deep learning approaches. We used de-identified treatment data of PLHIV collected through UgandaEMR - an electronic medical records (EMR) system implemented in over 1,000 health facilities across Uganda.

### 2.2 Dataset

The dataset used in this study consists of routinely collected, de-identified, longitudinal data of PLHIV accessing care and treatment services from ART facilities with the UgandaEMR system. We extracted records of 106,155 PLHIV with 2,071,479 clinical visits from 99 health facilities across 56 districts in Uganda. These facilities included 12 general hospitals, 8 Health Centre (HC) IIs, 53 HCIIIs, 19 HCIVs, 2 national referral hospitals, 2 regional referral hospitals, and 2 special clinics, which are facilities dedicated solely to providing HIV services (e.g., TASO clinics). The distribution reflects the range of health facility levels in Uganda’s HIV care system and indicates wide geographic coverage across the country. These data included PLHIV who were active in care between 2000 and 2023. It included demographic variables (age, sex, contact status), clinical-related variables World Health Organization (WHO) stage, tuberculosis (TB) status), treatment-related variables (treatment duration, adherence, antiretroviral therapy (ART) regimen), and structural variables (facility level, entry point).

### 2.3 Outcome Variable

The study outcome was defined using the PEPFAR definition of loss to follow-up (LTFU). A clinical visit was considered “attended” if a client came before or within 28 days of the next scheduled return visit date [20]. Clients within this window were considered to be retained in care at that particular time. A visit was labeled “LTFU” if a client’s clinical visit did not occur within 28 days of their scheduled return visit date.

### 2.4 Inclusion and Exclusion

The study included PLHIV with at least one clinical visit and well-defined data points across various variables, whose records were captured using UgandaEMR. PLHIV initiated into care between 2000 and 2023 were included in the study. We excluded clients with only one clinical visit because it was not possible to determine the outcome variable for those records. The outcome required at least two visits to establish whether a client returned within the expected timeframe. We also excluded clients initiated before the year 2000, and clinical visits missing the outcome variable due to missing the next visit date. Records missing a significant number of data points were also excluded. These records typically lacked nearly all key predictor variables and could not be used for modeling without introducing bias through imputation. Figure 1 shows the number of clients, clinical visits, and facilities included and excluded from the study.

**Figure 1.**
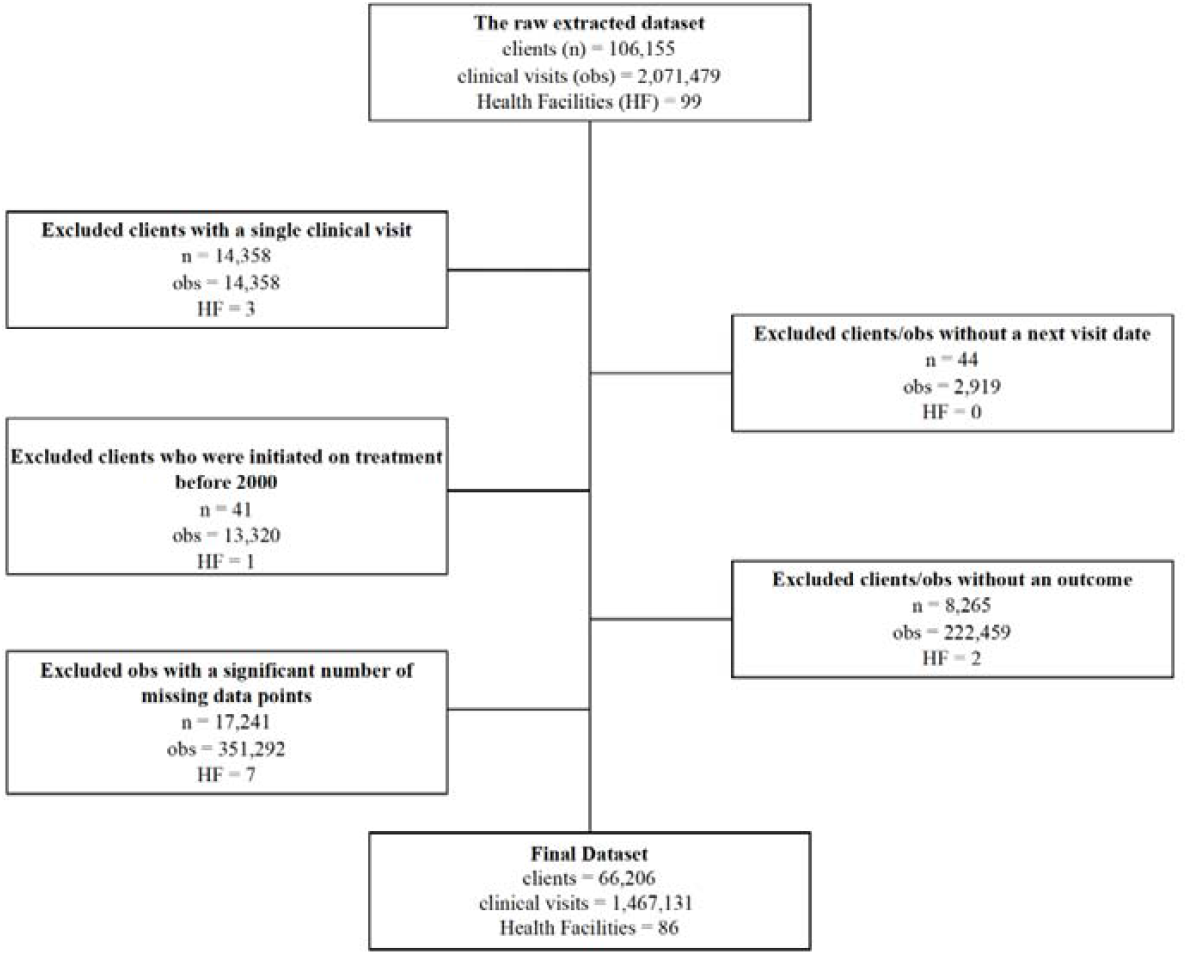
Flowchart of inclusion and exclusion criteria for clients and their associated clinical visits and health facilities

### 2.5 Ethical Consideration and Consent

This activity was reviewed by U.S. CDC, and was determined to be research not involving human subjects, and was conducted consistent with applicable federal law and CDC policy, under 45 C.F.R. part 46; 21 C.F.R. part 56; 42 U.S.C. §241(d), 5 U.S.C. §552a, 44 U.S.C. §3501, and it was also reviewed and approved by the Makerere University School of Public Health, Research and Ethics committee. The study involved a secondary analysis of de-identified data collected as part of routine clinical care. The Makerere University School of Public Health Research and Ethics Committee reviewed and approved the study, including a waiver of individual informed consent, as the data used were de-identified and posed minimal risk to participants.

### 2.6 Predictors and Feature Engineering

This study utilized 18 predictors, including both raw and desired/engineered variables, to develop more robust predictive models. The predictors included client demographics such as age and sex. Derived clinical visit patterns included the overall number of visits, the number of interruptions in treatment (IITs) a client has ever had, the number of IITs in the last 12 months, and the duration on treatment. Information on IITs was included to help the models capture historical treatment interruptions. All engineered features, including visit counts and treatment interruptions, were derived strictly from data available up to the prediction point to ensure that inputs reflected only information known at the time of prediction and to prevent data leakage. Treatment data variables included the current ART regimen, regimen line (categorized as first-line, second-line, or third-line based on national treatment guidelines), regimen changes, and the number of visits on the current regimen (i.e., the number of visits the patient has had while on their current regimen). Additionally, we included calculated variables such as the season, derived from the month of each return visit date. We categorized March, April, May, September, October, November, and December as the “rainy season” and January, February, June, July, and August as the “dry season” to investigate the effect of seasonality on clinical visit attendance, based on the National Meteorological Authority seasonal report. A full list and definitions of all predictors used in the models are provided in Appendix A.

### 2.7 Missing data

The level of completeness for the considered variables varied significantly, with some variables, such as age, sex, and entry point, having a 100% completeness rate, while others, like comorbidities, had a very low completeness rate of 1.5%. Variables with very low completeness, such as comorbidities, CD4 count, weight, and height, were eliminated from the analysis to avoid the bias that could be introduced by imputing these largely missing values. For the phone number variable, we transformed the data to indicate whether a contact was provided (yes) or not (no), replacing NA values with “no” under the assumption that the number was not provided.

For PLHIV missing an ART start date, we used the proxy of the first encounter date. Since most machine learning models except XGBoost cannot handle missing values directly, and because many of the variables were categorical, which could introduce bias if missing values were replaced with the majority class, we opted to remove all records with missing data [21]. This decision was supported by the negligible effect it had on the models’ performance. We also tested adding a missing category for selected variables, but this approach did not improve model performance and introduced sparsity, so it was not used.

### 2.8 Machine Learning

We explored several machine learning algorithms to compare their performance in predicting whether PLHIV will honor their scheduled appointment date. Models evaluated were: Decision Tree (DT), Random Forest (RF), AdaBoost, and Extreme Gradient Boosting (XGBoost). To preserve the temporal structure of the data and prevent information leakage, we applied a time-based split strategy at the encounter level. Specifically, all clinical visit records were first sorted in chronological order by visit date. The earliest 80% of encounters were assigned to the training set, and the most recent 20% to the testing set. To avoid information leakage across patient histories, we ensured that each PLHIV’s complete set of visits was assigned to only one partition [22]. If any of a patient’s visits fell into the test window, all of that patient’s visits were included in the test set. This ensured strict separation between training and test subjects while maintaining the real-world scenario of forecasting future outcomes using past data.

#### 2.8.1 Decision Trees

Decision Trees are a machine learning method that repeatedly splits the dataset according to a criterion that maximizes the separation of the data, resulting in a tree-like structure [23]. This study used a decision tree classifier and optimized its performance using a GridSearchCV approach. Several parameters were explored through a grid search, including the criterion (“gini” and “entropy”), splitter (“best” and “random”), max depth (None, 10, and 20), min samples split (2, 5, and 10), and min samples leaf (1, 5, and 10). The GridSearchCV was configured to use 10-fold cross-validation and optimize for accuracy. The optimal parameters identified were: criterion = ‘gini’, max depth = None, min samples leaf = 1, min samples split = 2, and splitter = ‘best’. This configuration was used to build the final Decision Tree models on the original dataset, as well as models using undersampling, oversampling, and SMOTE techniques to address class imbalance.

#### 2.8.2 Random Forest

Random Forest is an ensemble learning method that makes predictions by constructing multiple decision trees during training and outputting the class that is the mode of the classes or the mean prediction of the individual trees [24]. This method improves predictive accuracy and controls overfitting by averaging multiple decision trees, thereby reducing variance without significantly increasing bias. The Random Forest algorithm uses the principle of bagging, which combines the predictions of various base estimators built with different subsets of the dataset to improve generalizability [25].

In this study, the training procedure of the Random Forest model involved several steps to ensure optimal performance. We defined a parameter grid specifying the number of trees (100), maximum depth of each tree (10), minimum number of samples required to split a node (10), minimum number of samples required to be at a leaf node (4), and the number of features to consider for the best split (square root of the total number of features). The model training utilized grid search with 5-fold cross-validation to identify the best parameters. While an extensive grid search with multiple parameters was not performed due to the computational demands, the selected parameters were found to provide good performance.

#### 2.8.3 AdaBoost

AdaBoost, or Adaptive Boosting, is another ensemble learning technique that aims to improve the performance of weak classifiers by combining them into a strong classifier. In this study, we employed AdaBoost with a decision tree as the base classifier. AdaBoost works by sequentially fitting the base classifier to the data, where each subsequent model attempts to correct the errors made by the previous ones [26]. The algorithm assigns higher weights to misclassified instances, forcing the model to focus on the harder-to-classify samples. This iterative process continues until a specified number of classifiers are added or the performance no longer improves.

#### 2.8.4 Extreme Gradient Boosting

Extreme Gradient Boosting (XGBoost) is a highly efficient and scalable implementation of gradient boosting that has gained popularity for its performance in various machine learning competitions and practical applications [27]. The algorithm builds an ensemble of decision trees in a sequential manner, where each tree attempts to correct the errors made by the previous trees by minimizing a specified loss function using gradient descent optimization. XGBoost introduces several enhancements over traditional gradient boosting, such as regularization techniques to prevent overfitting, a tree learning algorithm for handling sparse data, and the ability to parallelize tree construction, significantly improving computational efficiency and scalability [27].

For this study, we utilized the XGBoost classifier with specific hyperparameters to optimize model performance. The parameters included a maximum tree depth of 6, a learning rate of 0.1, and 100 estimators. The model’s objective was set to binary logistic regression, suitable for binary classification tasks, and the booster type was set to use gradient-boosted trees. The XGBoost classifier was configured to use a single job for parallel processing, which banks on the algorithm’s ability to distribute computations across multiple processors. This approach not only accelerates the training process but also enhances the model’s ability to handle large datasets efficiently.

### 2.9 Bidirectional Encoder Representations from Transformers

We chose the BERT model due to its exceptional capability to handle sequential data and capture contextual information from both directions, which is critical in this study, where each HIV client has a longitudinal history of appointments. Traditional machine learning models, such as those discussed in Section 2.8, are designed to handle independent and identically distributed data, but struggle to capture the temporal dependencies within sequences of client visits [17]. In this work, we used a base pre-trained BERT model and fine-tuned it on the clinical dataset. BERT, on the other hand, leverages a transformer-based architecture that enables it to capture complex patterns and relationships within the sequential data by processing the entire sequence simultaneously [18]. This capability allows BERT to effectively learn patterns and trends from the sequence of past appointments, which is vital for accurately predicting future attendance. Studies have demonstrated the superiority of transformer-based models like BERT in modeling sequential data and capturing long-term dependencies compared to traditional methods [18, 19].

The model predicts the next appointment’s outcome based on the past appointments’ sequence, making it particularly useful for time-series prediction. Unlike traditional machine learning models that treat each clinical visit as independent, BERT uses past information to predict future outcomes. The prediction of the next appointment’s outcome was based on a sliding window approach to create sequences of a fixed length. Each sequence was used to predict the outcome of the subsequent appointments. A sequence length of the last 10 appointments was selected, balancing the need to capture sufficient recent historical data with model complexity. Studies have shown that recent behaviors are more predictive of future actions, and focusing on the last 10 appointments allows the model to leverage current trends and variations in a client’s attendance history [10, 28]. This length is also computationally efficient, enabling effective training and evaluation without excessive noise or overfitting.

At the core of the BERT model are six Transformer encoder layers, each comprising a multi-head self-attention mechanism and a position-wise fully connected feed-forward network [18]. The self-attention mechanism is crucial as it enables the model to weigh the importance of each past appointment in the sequence, thereby capturing relevant temporal patterns and dependencies [19]. To further refine the encoded information, the output from the Transformer encoder was passed through a fully connected layer with 512 units, followed by a dropout layer with a dropout rate of 0.3 to mitigate overfitting. The final classification layer maps the processed features to class probabilities corresponding to the predicted outcome. A dynamic calculation of the number of attention heads in the multi-head self-attention mechanism ensures compatibility with the input dimension, enhancing the model’s learning efficiency [18].

The Adam optimizer, known for its ability to handle sparse gradients, was used with a learning rate of 2×10^−5^. To adapt the learning rate dynamically during training, a StepLR scheduler was utilized. The training process was set for a maximum of 20 epochs, with early stopping implemented based on the validation loss to prevent overfitting.

### 2.10 Sampling

In normal circumstances, machine learning models are mainly evaluated using accuracy, precision, and recall score metrics [29]. However, in our study, we dealt with a class-imbalanced dataset, where the majority of clinical visits were attended (percent) compared to those missed (percent). This imbalance can make the standard metrics misleading, as the models may accurately predict clients who attended their appointments on time while poorly predicting missed appointments, which are critical for improving client retention in treatment.

To address this problem, we utilized sampling techniques including undersampling, oversampling, and the Synthetic Minority Over-sampling Technique (SMOTE). These methods were employed to assess their impact on the performance of the models compared to the results achieved when trained on the imbalanced dataset.

#### 2.10.1 Undersampling

Undersampling is a technique used to reduce the number of samples in the majority class to address the class imbalance in the dataset [30, 31]. This can be achieved either randomly, through Random under-sampling (RUS), or using statistical knowledge, through informed undersampling. For this study, the RUS method was used with a sampling strategy that undersamples the majority class to 90% of the minority class size. RUS reduces instances of the majority class by randomly deleting them from the dataset, thus creating a balanced dataset for both the majority and minority classes. This approach helps reduce the bias towards the majority class, allowing the machine learning models to perform better on the minority class.

#### 2.10.2 Oversampling

For oversampling, new samples are added to the minority class to balance the dataset. These methods can be categorized into random oversampling, where the existing minority samples are replicated to increase the size of a minority class, and synthetic oversampling, where artificial samples are generated for the minority class. These new samples add essential information to the minority class and prevent its instances from being misclassified [32]. Similar to the approach used in undersampling, we utilized a sampling strategy of 90%. This approach ensured that the models had sufficient data from the minority class, improving their ability to correctly classify minority class instances and reducing bias towards the majority class.

#### 2.10.3 SMOTE

SMOTE is a sampling method that generates synthetic samples within the minority class to address class imbalance[33]. SMOTE operates by identifying the minority class boundary samples, then producing a number of new samples between the boundary samples and the neighbors, and finally adding these synthetic samples to the original dataset[34]. In this study, the minority class in the training dataset was increased by over 90%. A significant number of synthetic instances of the minority class were generated and added to the dataset, enhancing the models’ ability to learn from a more balanced distribution and improving the accuracy of predictions for the minority class.

### 2.11 Model Validation

After training, the performance of all models was evaluated and compared to select the best-performing one for predicting missed appointment dates among HIV patients. The evaluation used confusion matrices to compute accuracy, precision, sensitivity, specificity, F1-score, and the area under the receiver operating characteristic (AUC-ROC), as these are the most widely used evaluation metrics for classification tasks in machine learning [45]. These metrics provided a comprehensive assessment of each model’s effectiveness in correctly identifying clients who were at risk of missing their appointments.

### 2.12 Implementation

The models were developed and evaluated using open-source Python libraries. Scikit-learn was used for traditional machine learning algorithms [35], while PyTorch was utilized for deep learning models, particularly the BERT model [18, 36]. Data cleaning and transformations were performed using Pandas [37], ensuring that the dataset was properly preprocessed for model training.

## 3 Results

The final dataset used in this study included 66,206 PLHIV with 1,479,121 clinical visits from 86 facilities. A descriptive statistical analysis was conducted to understand the demographic and detailed characterization of the clients (see Table 1). The dataset comprised 43,132 (65.1%) females with a median age of 36.0 years [IQR: 28.0 – 46.0 years], and 23,074 (34.9%) males with a median age of 41.0 years [IQR: 32.0 – 50.0 years]. Each PLHIV had an average of 22.34 clinical visits.

**Table 1:** Baseline characteristics of the study participants in Uganda, 2000-2023.

| Variable | Category | Frequency | Percentage |
| --- | --- | --- | --- |
| Client |  | 66,206 |  |
| Clinical Visits |  | 1,479,121 |  |
| Sex | Male | 23,074 | 34.9 |
|  | Female | 43,132 | 65.1 |
| Age | 0-13 | 2,447 | 3.7 |
|  | 15-20 | 2,889 | 4.4 |
|  | 21-35 | 22,796 | 34.4 |
|  | 36+ | 38,074 | 57.5 |
|  | Median (IQR) <sup>a</sup> | 38.0 (IQR: [29.0 - 47.0]) |  |
| Initiation year | 2000-2005 | 2,695 | 4.1 |
|  | 2006-2011 | 18,141 | 27.4 |
|  | 2012-2017 | 28,974 | 43.8 |
|  | 2018-2023 | 16,396 | 24.8 |
| WHO Stage <sup>b</sup> | I | 46,389 | 70.1 |
|  | II | 16,307 | 24.6 |
|  | III | 2,610 | 3.9 |
|  | IV | 900 | 1.4 |
| Advanced HIV <sup>b,c</sup> | Yes | 3,818 | 5.8 |
|  | No | 62,388 | 94.2 |
| Treatment Duration | < 6 months | 5,714 | 8.6 |
|  | 6 months -< 1 year | 3,870 | 5.8 |
|  | 1 -< 2 years | 6,198 | 9.4 |
|  | 2 -< 5 years | 13,194 | 19.9 |
|  | 5 years and above | 37,230 | 56.2 |
| TB Status <sup>b,d</sup> | Yes | 3,313 | 5.0 |
|  | No | 62,893 | 95.0 |
| Facility Entry Point | Exposed Infant | 53 | 0.1 |
|  | Outreach | 5,416 | 8.2 |
|  | KP Clinic | 240 | 0.4 |
|  | Medical Inpatient | 959 | 1.4 |
|  | Medical Outpatient | 33,212 | 50.2 |
|  | PMTCT | 4,632 | 7.0 |
|  | SMC | 15 | 0.0 |
|  | STI Outpatient | 185 | 0.3 |
|  | TB clinic | 143 | 0.2 |
|  | YCC Child Clinic | 182 | 0.3 |
|  | Others | 21,169 | 32.0 |
| Contact <sup>e</sup> | Yes | 20,226 | 30.6 |
|  | No | 45,980 | 96.4 |

<sup>a</sup> The median age and interquartile range of the clients, calculated at the 25<sup>th</sup> and 75<sup>th</sup> percentiles.
<sup>b</sup> Clients' status as of their last clinical visit.
<sup>c</sup> Advanced HIV: Clients marked with "Yes" have either a CD4 cell count <200 cells/mm<sup>3</sup>, are in WHO stage 3 or 4, or are children younger than five years old.
<sup>d</sup> TB status: Clients marked with "Yes" are either currently undergoing TB treatment or have been diagnosed with TB but are not on treatment.
<sup>e</sup> Contact: Clients with a "Yes" provided their phone contact numbers.

Overall, 158,266 (10.7%) of the clinical visits were missed, and 49,588 (74.9%) of PLHIV had at least one missed clinical visit. Figure 2 shows the trends in missed appointments from 2010 to 2023, focusing on years with more complete and reliable reporting. During this period, the proportion of missed appointments was relatively stable, ranging from a low of 9.1% to a high of 13.2%. This consistency highlights the persistent challenge of missed appointments and the continued need for effective interventions to improve adherence.

**Figure 2.**
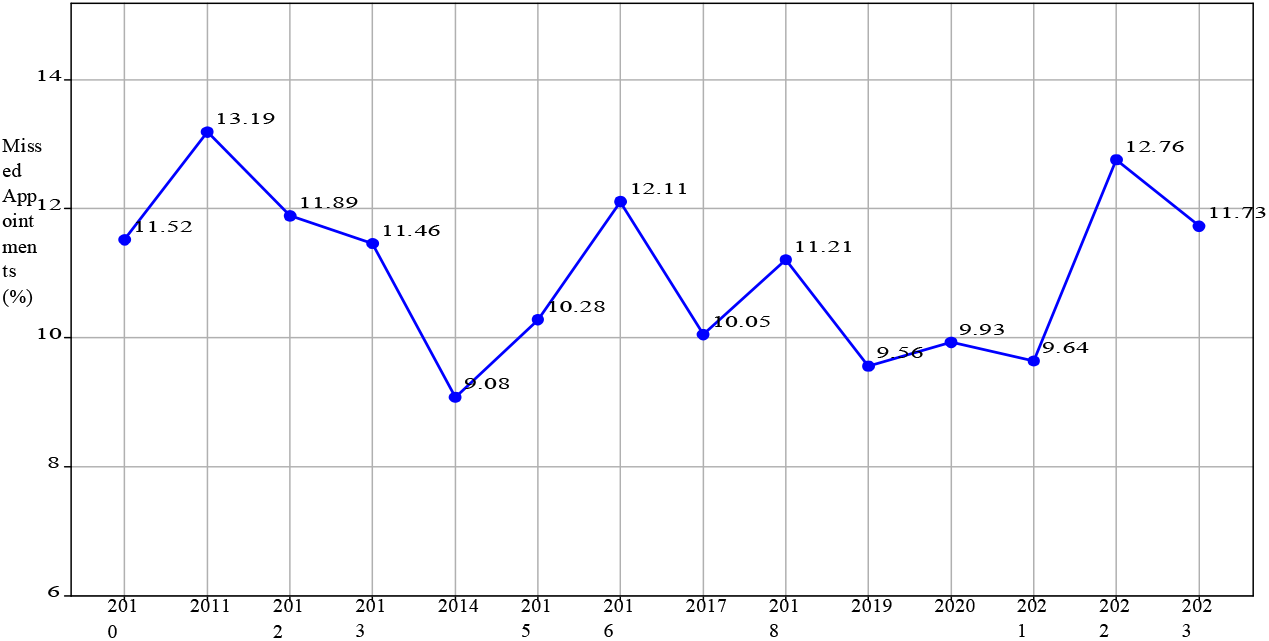
Trends in the proportion that missed appointments across different years from 2010 to 2023

### 3.1 Model Performance

The performance of different algorithms on the original dataset and with various sampling techniques is presented in Table 2. The Decision Tree classifier achieved an accuracy of 92.6% on the original dataset, with a precision, recall, and F1-score of 92.9%, 99.3%, and 96.0%, respectively. However, when undersampling was applied, the performance metrics dropped significantly, with accuracy at 79.7%, precision at 97.0%, recall at 79.8%, and F1-score at 87.5%. This reflects the expected trade-off where overall accuracy decreases, but the model improves its ability to correctly identify missed appointments, which was the primary outcome of interest. Oversampling and SMOTE improved the recall but had varying effects on other metrics, with SMOTE yielding a more balanced improvement across all metrics. Similar trends were observed with other models, such as Random Forest, AdaBoost, and XGBoost, which performed well on the original dataset but showed varied results when sampling techniques were applied.

**Table 2:**
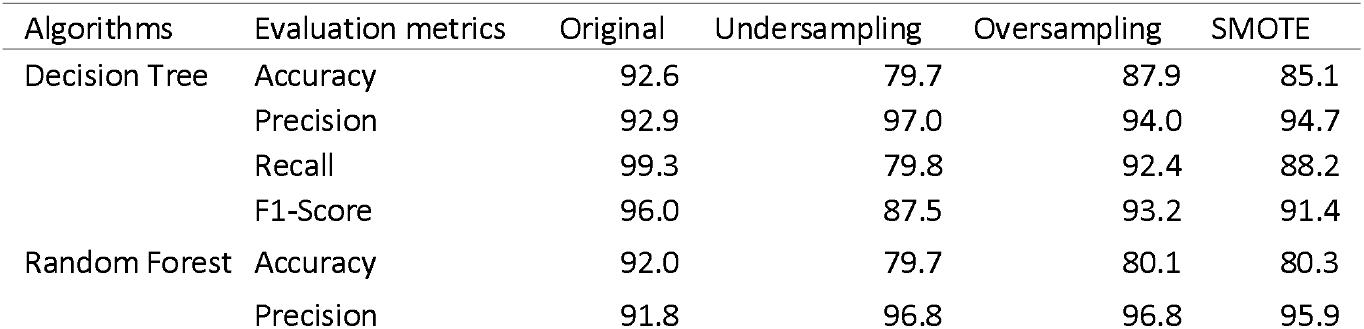
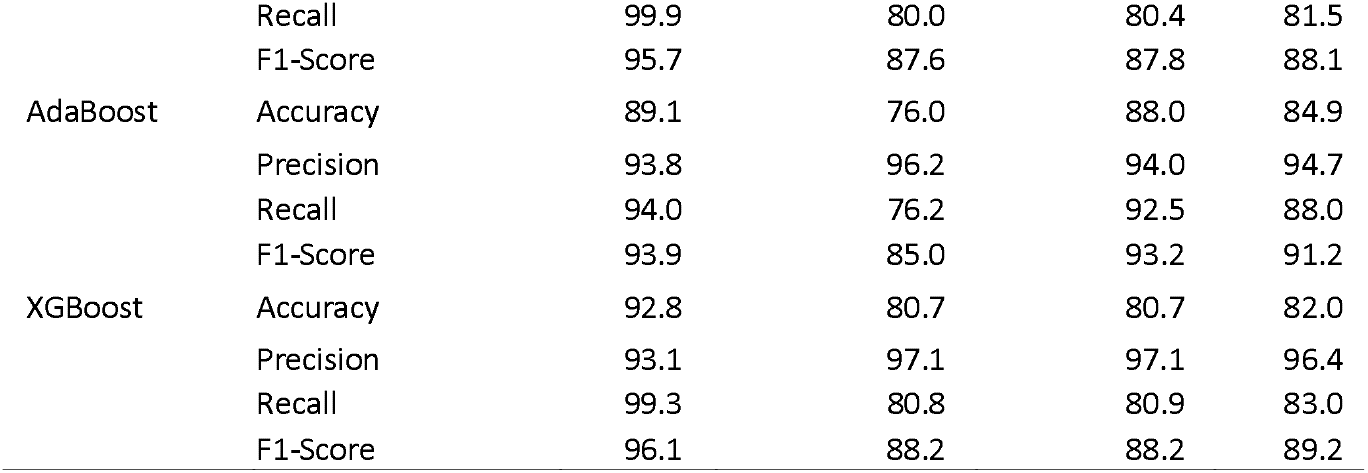
Performance metrics comparison of the algorithms trained on the original dataset and with different sampling techniques.

Although the models showed high performance on the original dataset, this was primarily due to the bias towards the majority class of clients who attended their treatment appointments. This apparent high performance on the original dataset can be misleading as it suggests the models are more effective than they truly are. To address this, we calculated the AUC scores for different models, revealing that those fitted using undersampling provided had more balanced performance in predicting both classes. As illustrated in Figure 3, the Random Forest and XGBoost models with undersampling had the highest AUC scores among the traditional machine learning models.

**Figure 3.**
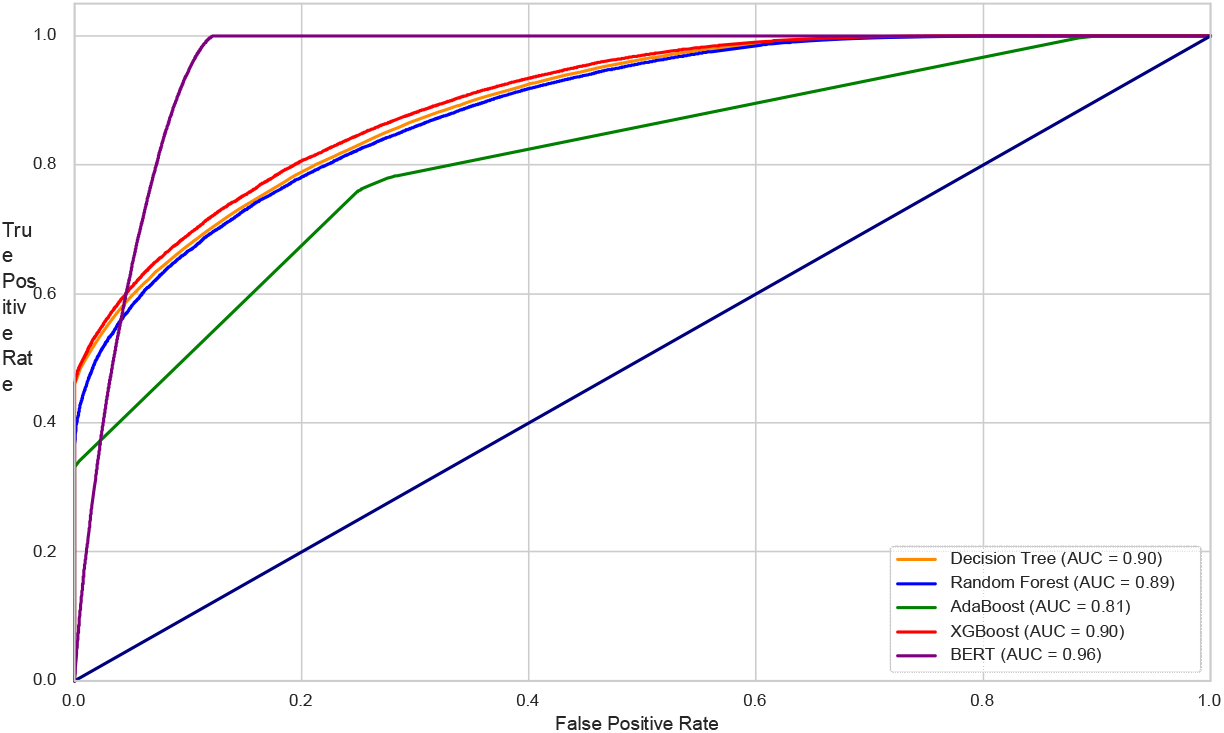
Model performance - AUC scores

When comparing traditional models with the BERT model, the latter demonstrated superior performance, with an AUC score of 0.96, indicating its effectiveness in distinguishing between classes and highlighting the superiority of transformers for sequential data. Figure 4 compares the performance of the best-performing traditional machine learning model (XGBoost with undersampling) and the BERT model across various metrics. The BERT model outperformed XGBoost in terms of accuracy (94.8% vs. 80.7%), precision (94.8% vs. 97.1%), recall (100.0% vs. 80.8%), and F1-score (94.2% vs. 88.2%).

**Figure 4.**
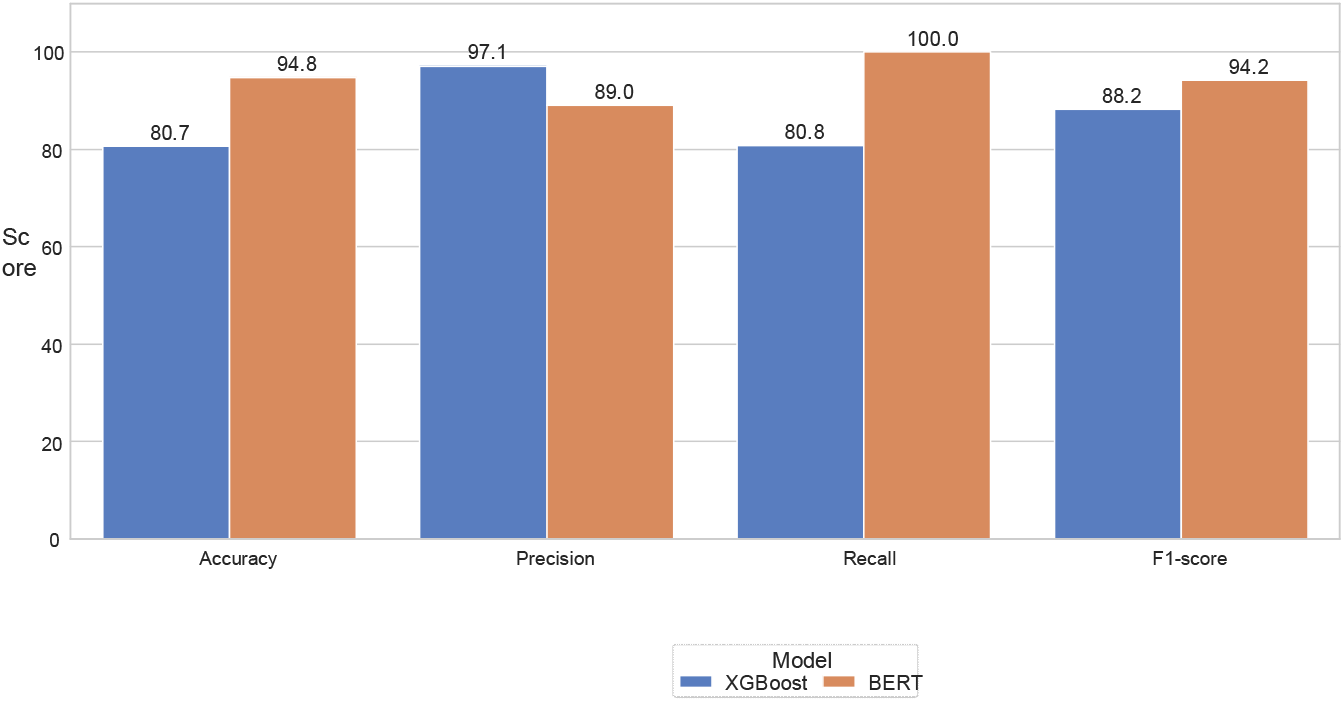
Overall model performance comparison between the best traditional machine learning model (XGBoost with Undersampling) and the BERT Model.

Further analysis of the model’s performance is provided in Table 3, which details the classification performance for each class (Attended and Missed). The BERT model achieved perfect precision (100%) and high recall (88.0%) for the ‘Attended’ class, and also excelled in classifying the ‘Missed’ class with a precision of 89.0% and recall of 100%, leading to F1-scores of 93.0% and 94.0%, respectively. In comparison, XGBoost, while showing high precision for the ‘Attended’ class (97.1%), had a lower recall (80.8%) and F1-score (88.2%). Other models exhibited varied performance, with Decision Tree and Random Forest showing relatively balanced results, while AdaBoost struggled more with the ‘Missed’ class.

**Table 3:**
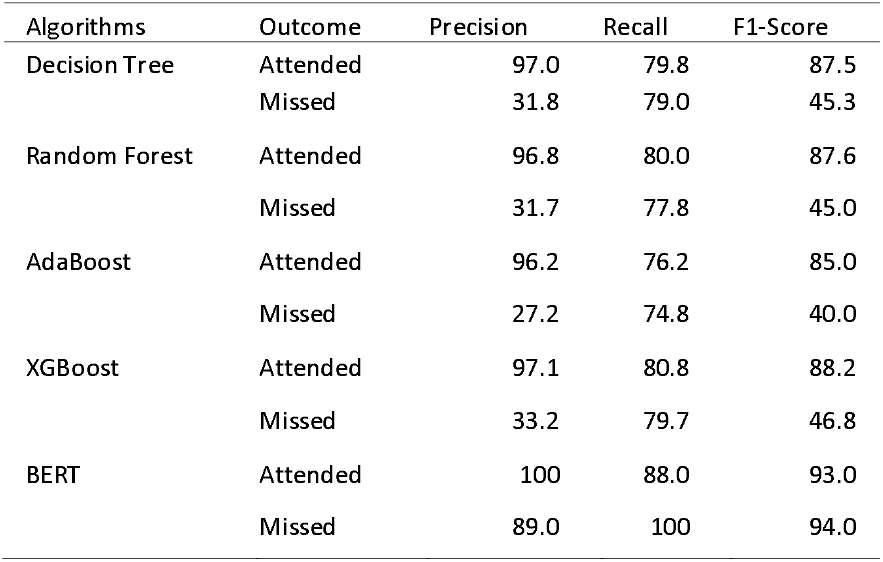
Performance of models in classifying each class.

| Algorithms | Outcome | Precision | Recall | F1-Score |
| --- | --- | --- | --- | --- |
| Decision Tree | Attended | 97.0 | 79.8 | 87.5 |
|  | Missed | 31.8 | 79.0 | 45.3 |
| Random Forest | Attended | 96.8 | 80.0 | 87.6 |
|  | Missed | 31.7 | 77.8 | 45.0 |
| AdaBoost | Attended | 96.2 | 76.2 | 85.0 |
|  | Missed | 27.2 | 74.8 | 40.0 |
| XGBoost | Attended | 97.1 | 80.8 | 88.2 |
|  | Missed | 33.2 | 79.7 | 46.8 |
| BERT | Attended | 100 | 88.0 | 93.0 |
|  | Missed | 89.0 | 100 | 94.0 |

Figure 5 illustrates the feature importance for predicting PLHIV likelihood of attending their next clinical visit. The features are ranked by their relative importance, highlighting the variables that contribute most significantly to the model’s predictions. The top three predictors were: PLHIV history of interruptions in treatment (IIT), the total number of visits, and recent treatment interruptions are the most influential factors. These were followed by overall duration of treatment, the year of treatment initiation, and the number of visits under the current ART regimen are also significant predictors. In contrast, variables such as “sex”, “TB”, and “season”, were shown to have minimal impact on the predictions.

**Figure 5.**
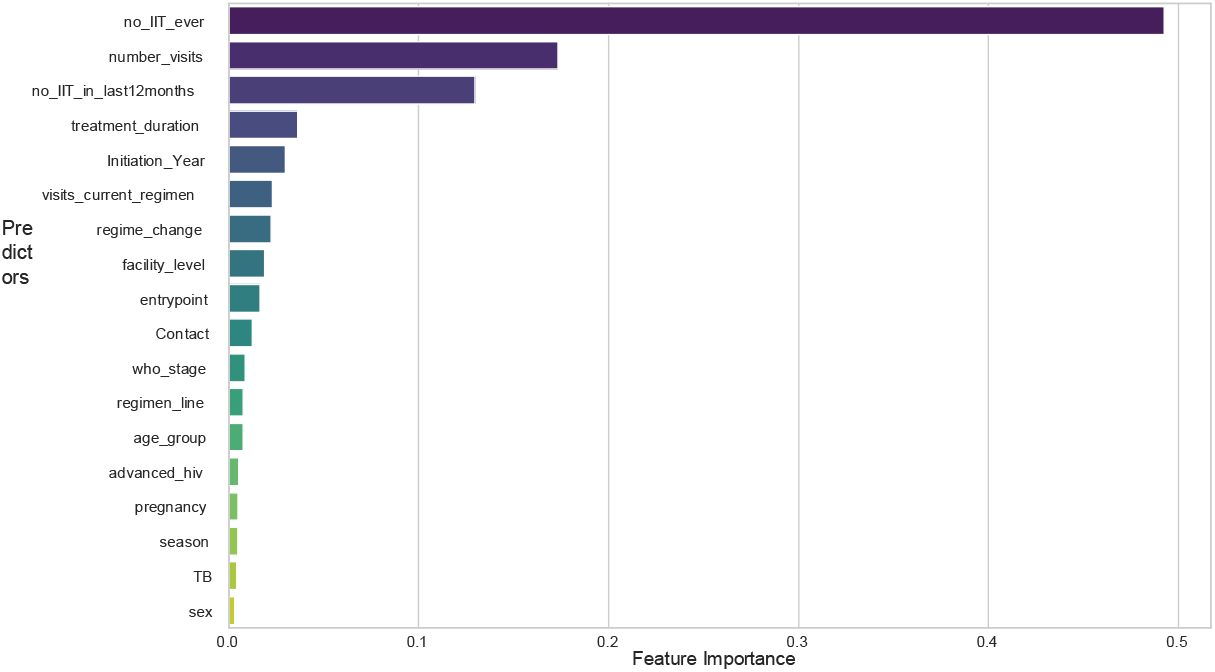
Feature importance of variables ranked by their contribution to predicting the likelihood of a client attending their next clinical visit.

## 4 Discussion

The results from our comparative analysis show that the BERT model performed better than the traditional machine learning models in predicting missed appointment treatment dates among HIV patients using longitudinal clinical data. The BERT model achieved an AUC score of 0.96, higher than the scores obtained by the Decision Tree, Random Forest, AdaBoost, and XGBoost models. This superior performance can be attributed to BERT’s ability to capture complex patterns and dependencies within sequential data, an important factor in time-series prediction tasks such as appointment attendance [38]. Traditional models like Decision Trees and Random Forests treat each instance independently, limiting their ability to leverage temporal information [16, 17]. In contrast, BERT’s transformer architecture allows it to consider the sequence of appointments holistically, enhancing its predictive accuracy and robustness [18, 19]. These results demonstrate that when machine learning and deep learning approaches are properly applied to routinely collected clinical and demographic data, they can correctly predict appointment attendance with high accuracy.

The BERT model exhibited very high precision when predicting PLHIV likely to honor their treatment schedules, indicating it is almost always correct in its positive predictions. Its recall was slightly lower, meaning some actual attended visits were missed, but the high F1-score showed a strong balance between the two. For missed appointments, the BERT model had high recall, correctly identifying all PLHIV likely to miss their treatment appointment schedules, though its precision was slightly lower, indicating some false positives. These results align with findings from Ramachandran et al. [11], who used predictive models in a U.S. HIV clinic and achieved a positive predictive value of 34.6% for the top 10% high-risk clients, and from Maskew et al. [10], who reported an AUC of 0.69 using random forest models in South Africa. Compared to these, BERT achieved higher precision (89.0% for missed visits), recall (100%), and AUC (0.96), suggesting that transformer-based models offer superior performance in handling sequential clinical data. This demonstrates the advantage of using temporal models like BERT over traditional approaches in predicting both attendance and risk of disengagement.

The impact of various sampling techniques on the performance of traditional machine learning models showed that undersampling generally resulted in a performance drop across all metrics for the Decision Tree, Random Forest, AdaBoost, and XGBoost models due to the loss of valuable information when the majority class is reduced. However, undersampling provided a balance in performance for the two classes, evident from the improved AUC scores for Random Forest and XGBoost. Conversely, oversampling and SMOTE helped maintain more stable performance by reducing bias towards the majority class while retaining sufficient samples for training. These sampling techniques effectively addressed the imbalance problem, yielding better prediction outcomes for both classes. The algorithms achieved high accuracy scores when trained on the original imbalanced dataset, which can be misleading due to the imbalance in outcome classes. Correctly predicting PLHIV who are likely to miss their appointments is important for improving retention in care. Therefore, although high performance on an imbalanced dataset may appear impressive, it does not necessarily reflect true predictive accuracy. The use of sampling techniques significantly improved the classification accuracy for both classes. Similar findings were reported by Cai et al. [39], who applied SMOTE to improve the performance of models predicting retention in HIV care, and Bero et al. [40], who found that oversampling improved the sensitivity of models identifying clients at risk of dropping out of PrEP programs. Based on our findings, if computational resources and expertise are limited, the XGBoost model with undersampling provides the most balanced performance among traditional machine learning models evaluated.

The feature importance analysis highlighted the critical role of historical treatment patterns in predicting adherence to appointments. In particular, interruptions in treatment and visit frequency emerged as dominant factors, showing that past engagement strongly influences future outcomes. These findings suggest that monitoring adherence history and visit consistency is central to identifying clients at risk of missing appointments. While demographic characteristics such as sex, age group, and seasonal factors contributed less, they still provide additional context for risk stratification. Overall, these results align with other studies showing that patient behavior and treatment history are stronger predictors of retention than demographic variables, emphasizing the value of personalized treatment support and closer monitoring of continuity in care [10,11,40].

### 4.1 Limitations and Future Work

The results of this study should be interpreted with some limitations in mind. First, the dataset had substantial missing or undefined values, resulting in the exclusion of 39,949 (37.6%) clients and 604,348 (29.2%) clinical visits. This reduction in the analytic sample may introduce selection bias and limit the extent to which the findings can be generalized to the broader population of PLHIV in care. Additionally, factors influencing PLHIV clinical attendance, such as education level and community contextual factors, were not routinely captured in the UgandaEMR. Also, mislabeling of the outcome was another limitation, as health facilities sometimes did not update transfer-in and out statuses in the EMR promptly. This may have led to PLHIV who were transferred to another facility being incorrectly labeled as having missed a visit.

We did not explicitly account for the hierarchical structure of patients nested within health facilities. Although facility-level variables were included, unmeasured differences across sites, such as urban versus rural setting, staffing levels, or resource availability, may have introduced clustering effects not captured in our models. Future work could address this using multilevel or mixed-effects modeling.

The interpretability of complex models like BERT remains a challenge, as it is difficult to explain how and why these models arrive at their predictions, making them somewhat of a black box [41]. Finally, the BERT model does not account for irregular time intervals between clinical visits. It processes sequences in fixed order, assessing equal spacing between events, which may obscure time-dependent relations. Future work will focus on developing models that are both more interpretable and time-aware, incorporating approaches such as Time-BERT [42], transformer models with temporal decay [43], or recurrent models like GRU-D [44] to better capture irregular clinical patterns while improving the transparency of model predictions.

### 4.2 Study Contributions

Despite the mentioned limitations, our study offers several contributions. It is among the few to evaluate transformer models like BERT on clinical HIV data. The BERT model outperformed all traditional models regardless of the sampling technique used, indicating its superior ability to manage the complexities of the data. This analysis provides a comprehensive comparison between traditional machine learning models and deep learning approaches, demonstrating how sampling techniques impact the performance of these algorithms. This comparison serves as a foundation for other researchers interested in similar studies. The study also responds to the call by the Government of Uganda through the Ministry of Health to integrate cutting-edge technologies like artificial intelligence into clinical decision-support systems to improve care and treatment.

Lastly, and most importantly, the developed models will be integrated into the UgandaEMR to offer a more proactive treatment implementation. This integration will help clinicians and HIV caregivers identify PLHIV at risk of missing their treatment schedules, thereby improving retention in HIV care, enhancing viral load suppression, and closing the gap to achieve the 95-95-95 UNAIDS targets.

## 5 Conclusion

This study highlights the potential of advanced machine learning and deep learning models, particularly transformer-based architectures like BERT, in predicting missed appointment dates among HIV patients. Our implementation framework enables clinicians and HIV caregivers to identify clients at the highest risk of dropping out of care and develop targeted interventions to improve retention. The results show that BERT significantly outperforms traditional machine learning models. This superior performance is attributed to BERT’s ability to effectively capture and utilize complex temporal patterns within sequential data, which is vital for time-series prediction tasks such as appointment adherence.

Even with various sampling techniques applied to traditional models to address class imbalance, they could not match the accuracy of the BERT model. These findings indicate that while techniques such as undersampling, oversampling, and SMOTE can improve the performance of traditional models, advanced architectures like BERT are better suited for handling complex, sequential clinical datasets. This insight is valuable for guiding future research and implementations aimed at optimizing HIV patient care through predictive modeling and proactive programmatic strategies.

## Funding

This project has been supported by the President’s Emergency Plan for AIDS Relief (PEPFAR) through the Centers for Disease Control and Prevention under the terms of the 6NU2GGH0022280-04-00. The findings and conclusions in this report are those of the authors and do not necessarily represent the official position of the Centers for Disease Control and Prevention.

## Conflict of interest

The authors report no conflicts of interest.

## Data Availability

Due to the sensitive nature of the research, which involves client-level HIV data, the dataset is not publicly available.

## Code Availability

The code used for data preprocessing, transformation, and modeling can be made available upon request. Interested parties should contact the corresponding author to obtain access.

## Author contribution

A.M. contributed to conceptualization, data extraction, formal analysis, methodology, and writing— original draft.

A.G.F. contributed to formal analysis, methodology, and writing—review and editing. E.S. and K.M. contributed to conceptualization and writing—review and editing.

S.S. contributed to data extraction and writing—review and editing.

A.N., E.A., S.M., M.K., J.M., P.K.,N.M, P.M., C.A. contributed to supervision and writing—review and editing.

## Abbreviations

ART: Antiretroviral Therapy
AUC-ROC: Area Under the Receiver Operating Characteristic Curve
BERT: Bidirectional Encoder Representations from Transformers
EMR: Electronic Medical Records
IIT: Interruption in Treatment
LTFU: Loss to follow-up
PLHIV: People Living with HIV
SMOTE: Synthetic Minority Over-sampling Technique UNAIDS Joint United Nations Programme on HIV/AIDS
XGBoost: Extreme Gradient Boosting

## Appendix A

List of all predictor variables used in the models, along with their definitions to support reproducibility and clarity of interpretation.

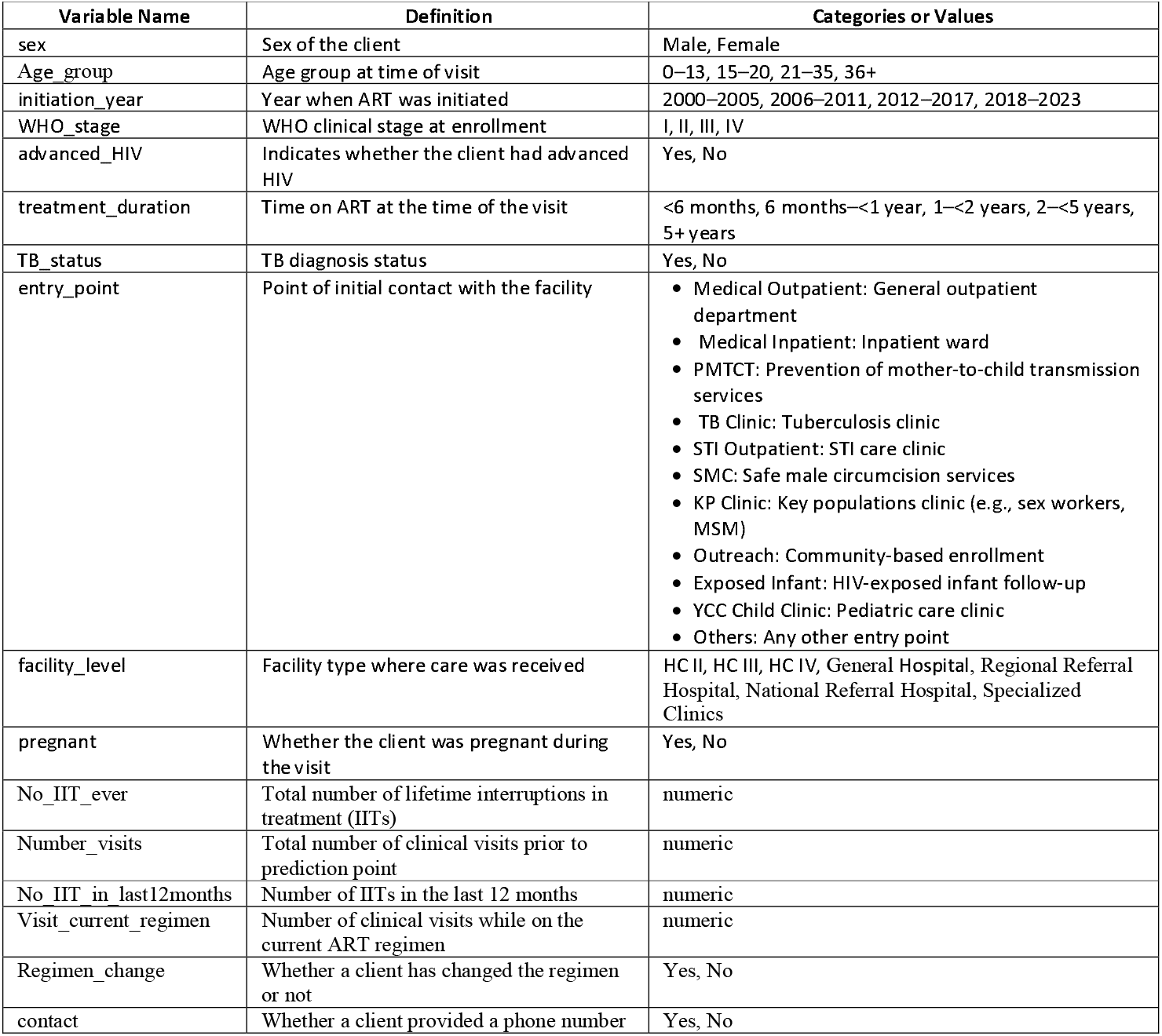
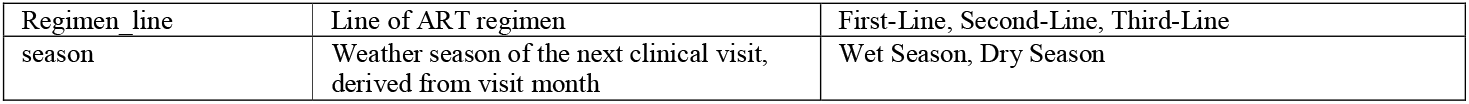

| Variable Name | Definition | Categories or Values |
| --- | --- | --- |
| sex | Sex of the client | Male, Female |
| Age_group | Age group at time of visit | 0–13, 15–20, 21–35, 36+ |
| initiation_year | Year when ART was initiated | 2000–2005, 2006–2011, 2012–2017, 2018–2023 |
| WHO_stage | WHO clinical stage at enrollment | I, II, III, IV |
| advanced_HIV | Indicates whether the client had advanced HIV | Yes, No |
| treatment_duration | Time on ART at the time of the visit | <6 months, 6 months–<1 year, 1–<2 years, 2–<5 years, 5+ years |
| TB_status | TB diagnosis status | Yes, No |
| entry_point | Point of initial contact with the facility | Medical Outpatient: General outpatient department<br>Medical Inpatient: Inpatient ward<br>PMTCT: Prevention of mother-to-child transmission services<br>TB Clinic: Tuberculosis clinic<br>STI Outpatient: STI care clinic<br>SMC: Safe male circumcision services<br>KP Clinic: Key populations clinic (e.g., sex workers, MSM)<br>Outreach: Community-based enrollment<br>Exposed Infant: HIV-exposed infant follow-up<br>YCC Child Clinic: Pediatric care clinic<br>Others: Any other entry point |
| facility_level | Facility type where care was received | HC II, HC III, HC IV, General Hospital, Regional Referral Hospital, National Referral Hospital, Specialized Clinics |
| pregnant | Whether the client was pregnant during the visit | Yes, No |
| No_IIT_ever | Total number of lifetime interruptions in treatment (IITs) | numeric |
| Number_visits | Total number of clinical visits prior to prediction point | numeric |
| No_IIT_in_last12months | Number of IITs in the last 12 months | numeric |
| Visit_current_regimen | Number of clinical visits while on the current ART regimen | numeric |
| Regimen_change | Whether a client has changed the regimen or not | Yes, No |
| contact | Whether a client provided a phone number | Yes, No |
| Regimen_line | Line of ART regimen | First-Line, Second-Line, Third-Line |
| season | Weather season of the next clinical visit,<br>derived from visit month | Wet Season, Dry Season |

## Notes

### Competing Interest Statement

The authors have declared no competing interest.

